# Characterizing HIV and STIs among Transgender Female Sex Workers: A Longitudinal Analysis

**DOI:** 10.1101/2020.03.12.20035139

**Authors:** Tonia Poteat, Rebecca Hamilton White, Katherine H.A. Footer, Ju Nyeong Park, Noya Galai, Steve Huettner, Brad E. Silberzahn, Sean T. Allen, Jennifer Glick, S. Wilson Beckham, Charlotte A. Gaydos, Susan G. Sherman

**Author notes:** Corresponding author Tonia Poteat, PhD, MPH, PA-C, University of North Carolina at Chapel Hill, 333 S. Columbia Street, #CB 7240, Chapel Hill, NC 272599-7240.

## Abstract

**Objectives:** Though highly vulnerable to HIV and STIs, transgender female sex workers (TFSW) are understudied in the U.S. HIV and STI response. This study examined the correlates of laboratory-confirmed STIs among a cohort of 62 TFSW followed over the course of one year and explored associations between specimen site and self-reported engagement in insertive and receptive anal intercourse.

**Methods:** Participants completed an interviewer-administered computer assisted personal interview at baseline, 3-, 6-, 9-, and 12-month visits where self-administered anal swabs and urine samples for gonorrhea, chlamydia, and trichomonas were also collected. HIV testing was conducted at baseline, 6-, and 12-month visits.

**Results:** Baseline HIV prevalence was 40.3% with no HIV seroconversions over follow-up. Baseline prevalence of gonorrhea, chlamydia, and trichomonas was 9.7%, 17.7%, and 14.5%, respectively. In the multivariable regression modeling, recent arrest was significantly associated with testing positive for any STI (aRR 1.77; 95% CI: 1.10-2.84). Insertive anal sex with clients was associated with increased risk of testing positive for an STI via urine specimen (RR 3.48; 95% CI: 1.14-10.62), while receptive anal sex was not significantly associated with specimen site.

**Conclusion:** Our findings confirm a high prevalence of STIs among TFSW and highlight the importance of addressing structural drivers such as criminal justice involvement as well as the need to ensure screening for STIs at all anatomical sites regardless of self-reported sites of potential exposure. More research is needed to better understand HIV and STI vulnerabilities and appropriate interventions for TFSW in the U.S.

**KEY MESSAGES:**

- Transgender female sex workers in the U.S. face substantial vulnerability to HIV and STIs, and interventions to reduce their risk for HIV and STI acquisition are needed.
- Engagement with criminal justice significantly increases the risk for STIs among transgender women engaged in street-based sex work, suggesting the need to address factors that lead to disproportionate arrest and incarceration of transgender women.
- Screening for STIs among transgender women should include all potential sites of exposure, regardless of reported sexual positioning.

## INTRODUCTION

Sex workers are highly vulnerable to HIV and sexually transmitted infections (STIs), globally.^1^ Yet, they remain understudied in the HIV/STI response, particularly in the United States (U.S.).^2^ Existing research with sex workers in the U.S. primarily focuses on cisgender female sex workers (CFSW), often to the exclusion of other vulnerable populations who engage in sex work.^3^ Largely due to pervasive employment discrimination, a substantial proportion of transgender women engage in sex work.^4^ A recent meta-analysis by researchers at the U.S. Centers for Disease Control and Prevention synthesized data from 29 studies to estimate that approximately 38% (95% CI: 29.0, 47.7) of transgender women in the U.S. have engaged in sex work (range 3.7-82%).^5^

Complex structural factors drive the high prevalence of HIV and STIs among transgender female sex workers (TFSW), including discriminatory application of anti-prostitution laws, widespread gender-based violence, and poor access to gender-affirming healthcare.^6^ In a study of racially and ethnically diverse transgender women in Northern California with a history of sex work, Nemoto and colleagues found a self-reported HIV prevalence of 30%.^7^ Twenty-six percent of this study’s participants also reported having had an STI in the past 12 months. Similar to data from CFSW, TFSW were more likely to engage in condomless receptive sex with primary partners (55%) than clients (23%). Nemoto found that condomless sex with primary partners was significantly associated with depressive symptoms and lower levels of self-esteem. Participants with higher levels of experienced transphobia, economic pressure, and need for social support; or who had lower self-esteem and self-efficacy were more likely to have engaged in condomless sex with clients.

In one of the few incidence studies among TFSW, Nuttbrock and Hwahng^8^ found a cumulative HIV incidence of 2.8% across their 3-year study with racially and ethnically diverse transgender women in New York. For years 1-3 of the study, new infections of syphilis were 3.6%, 1.1%, and 1.8%, respectively. Gonorrhea declined from 4.2% during year 1 to 0.0% during year 3; and chlamydia declined from 4.5% during year 1 to 1.1% during year 3. Altogether, the incidence rate for HIV and other STIs was 17.4% over the course of the study. In the same study, Black and Latina participants were more likely to report condomless receptive anal intercourse and, as a result, were more likely to acquire HIV or STIs than White participants.

The aforementioned studies provide important context for the high HIV and STIs risk among TFSW. However, these studies are limited by either lack of laboratory confirmation^7^ or single-site STI testing.^8^ For populations at high risk for STIs, current guidelines recommend screening in all sites of exposure which may include oral, anal, urethral and/or vaginal.^9^ In this study, we examined the correlates of laboratory-confirmed STIs at urethral and anal sites of exposure among a cohort of TFSW over one year. In order to assess potential drivers of HIV/STIs in TFSW, we tested for associations between STI results and baseline demographics, age of sex work entry, number of clients in the past 3 months, risk behavior (e.g., condomless sex), and structural factors (e.g., recent arrest). Additionally, we conducted a sub-analysis by STI specimen site (anal or urethral) to explore associations between specimen site and recent reported engagement in insertive and receptive anal intercourse.

## METHODS

### Study Design

The Sex workers And Police Promoting Health In Risky Environments (SAPPHIRE) study was a prospective cohort of cisgender and transgender women involved in street-based sex work, recruited between April 2016 and August 2017 and followed longitudinally for a year with visits at 3, 6, 9, and 12 months after their baseline visit.^10^ CFSW were recruited via targeted sampling across 14 zones in Baltimore City, MD, while TFSW were primarily recruited in one location identified through focus groups and ethnographic data. Detailed information about the study and recruitment strategies is available elsewhere.^11^ This analysis has been limited to the TFSW participants (n=62).

### Study participants

Eligibility criteria were: (1) age ≥ 15 years; (2) sold or traded oral, vaginal, or anal sex “for money or things like food, drugs or favors”; (3) picked up clients on the street or at public places ≥ 3 times in the past 3 months; (4) willing to undergo HIV and STI testing. Exclusion criterion was identifying as male or a man. Participants assigned male sex at birth who identified as women were included in the TFSW cohort.

### Study Procedures

Participants completed a 50-minute interviewer-administered computer assisted personal interview at each study visit. Biological specimens were also collected for HIV and STI testing. Participants who moved out of Baltimore City during follow-up remained eligible to participate. Surveys were administered via telephone if they moved more than a one-hour drive from the city, and HIV/STI testing was not done for participants who completed interviews by telephone. Study staff attempted to reach all participants who had a positive STI test result to encourage treatment and provide referrals. The study was reviewed and approved by the Johns Hopkins Bloomberg School of Public Health Institutional Review Board.

### Biological Measures

***HIV testing*** was conducted at baseline, 6-, and 12-month follow-up visits using OraQuick^©^ Advanced Rapid HIV-1/2 test kit (Orasure Technologies, Bethlehem, PA, USA). Results, counseling, and referrals for health and social services were delivered at the end of the interview. Self-administered anal swabs (Aptima swabs; Hologic, Inc., San Diego, CA, USA) and urine samples were collected at each study visit for ***STI testing***. Samples were analyzed using nucleic acid amplification tests (NAAT) for *Neisseria gonorrhoeae* (NG), *Chlamydia trachomatis* (CT), and *Trichomonas vaginalis* (TV) at the International Sexually Transmitted Diseases, Respiratory, and Emerging Diseases Research Laboratory at Johns Hopkins School of Medicine. Positive test results and participant contact information were sent to the Baltimore City Health Department and assigned to a Disease Intervention Specialist for linkage to treatment. Participants were also notified of their STI test results at their next study visit. TFSW who participated in follow-up interviews via telephone were not required to submit specimens for biological testing. Participants who tested positive for either NG, CT, or TV were categorized as positive for an STI.

### Psychosocial and Behavioral Measures

Structured interview measures were developed from previous research, existing scales, and engagement with the Community Advisory Board. We collected information on age, race, ethnicity, sex work, housing, finances, criminal justice involvement, sexual and drug use behaviors, health service access, and mental health. Clients were defined as anyone with whom participants had oral, vaginal, or anal sex for money or goods (e.g. food, drugs, or favors). Intimate partners (IP) were defined as non-paying, regular sexual partners. Sex work history measures included age first sold sex (street-based or otherwise), number of clients in the past 3 months dichotomized at the median (19 clients), and ever meeting clients online. Structural factors included homelessness (measured in the past 3 months), criminal justice history, unemployment at time of survey administration, and food insecurity (measured in the past 12 months). At baseline, arrest was measured in the past 12 months; over follow-up visits, arrest was measured in the past 3 months. We also explored physical or sexual violence separately over the past 3 months by perpetrator type using an adapted version of the Revised Conflicts Tactic Scale.^12^ Lifetime violence was a composite measure (range 0 to 4) of the types of violence TFSW experienced in their lifetime, including childhood physical or sexual abuse and physical or sexual abuse as an adult. Condom coercion was defined as condom removal or refusal in the past 3 months.^13^ Psychological factors addressed include depressive symptoms and posttraumatic stress disorder (PTSD). Depressive symptoms were assessed using the Center of Epidemiologic Studies Depression Scale-10 with a possible range of 0-30 and a cutoff of score of 10 indicative of clinically significant symptoms of depression.^14^ We used the PCL-5 scale to assess for symptoms of PTSD (possible range: 0-80) with a cutoff score of 33 to indicate a provisional PTSD diagnosis.^15^ Risk behaviors included the participant being high or drunk during sex in the past 3 months, which was dichotomized to always or often versus sometimes, rarely or never; engaging in condomless sex with clients and IPs; types of sex had with clients or IPs (insertive and receptive anal sex); substance use, which excludes marijuana use and includes any injection or non-injection drug use as only a few women reported ever injecting drugs (n=3); and having sex with clients in a public location in the past 3 months, which includes street, alley, car, abandoned building, park, or public bathroom. The full list of measures is provided in Table 1.

**Table 1.**
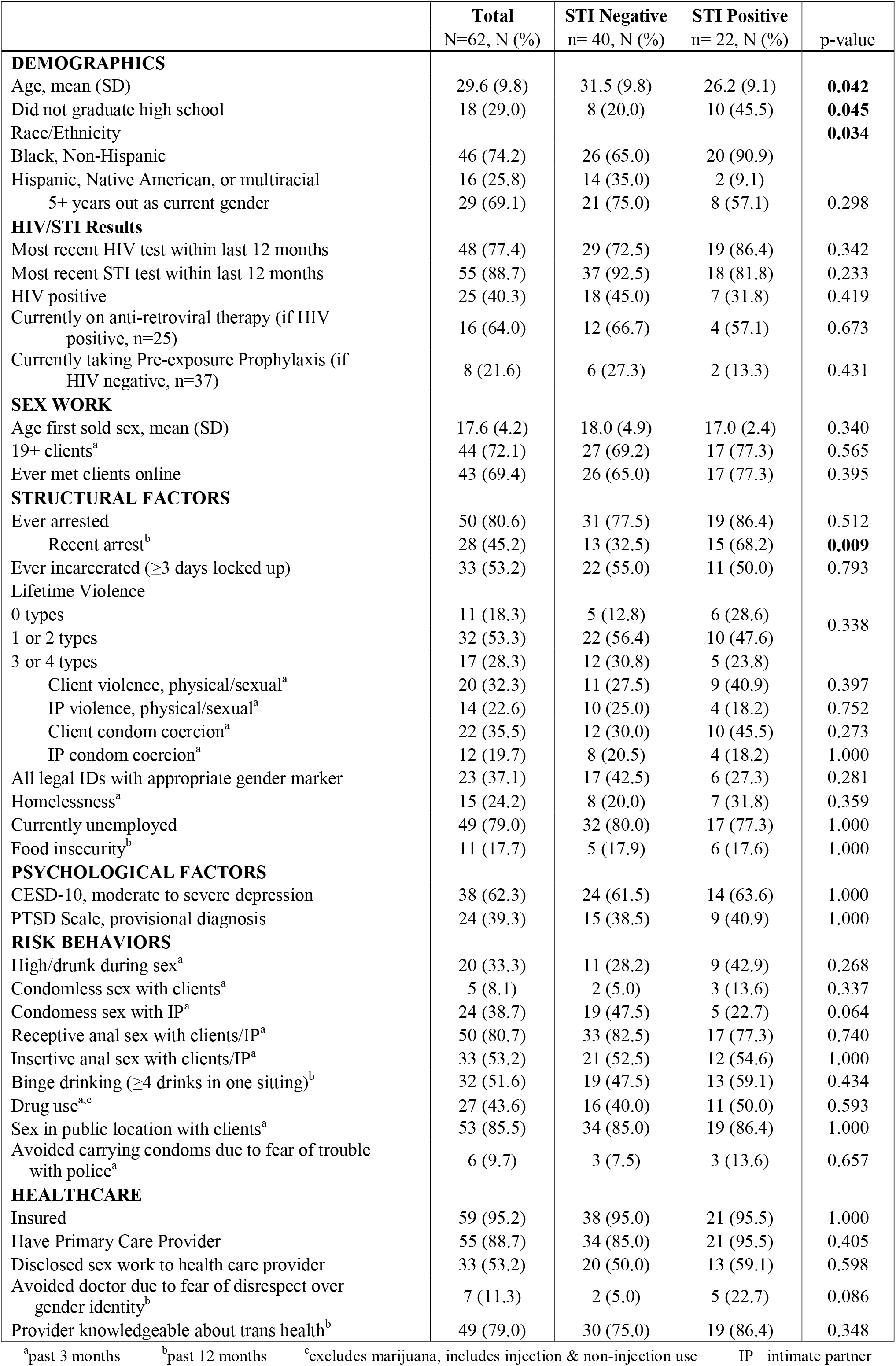
Baseline participant characteristics by positive at baseline versus negative at baseline for an STI (GC, CT, TV).

### Statistical Analyses

We compared TFSW who tested positive for any STI (NG, CT, or TV) from any site (urethral or anal) at baseline to TFSW who tested negative for all three STIs using Fisher’s exact tests and independent sample t-tests. Using longitudinal data from all study visits, we applied univariate log binomial regressions using General Estimating Equations (GEE) with exchangeable correlation structure to examine correlates of repeated STI episodes. An STI episode was defined dichotomously at each study visit as testing positive for any STI (NG, CT, or TV) by any specimen type (anal or urine) versus no positive results. We systematically explored all variables in Table 1 with univariate models, showing the most relevant measures in table 3. Covariates with *P*<0.20 at the bivariate level were included in the multivariable model examining predictors of any STI. Model results presented are risk ratio (RR) of an STI, 95% confidence intervals, and *p*-values with the threshold of statistical significance held at *p*<0.05. We conducted a univariate sub-analysis by STI specimen site (anal or urine sample) and recent reported receptive or insertive anal sex using log binomial models with GEE and exchangeable correlation structure. Statistical analyses were conducted using Stata/SE 15.1 (StataCorp, College Station, TX, USA).

**Table 3.**
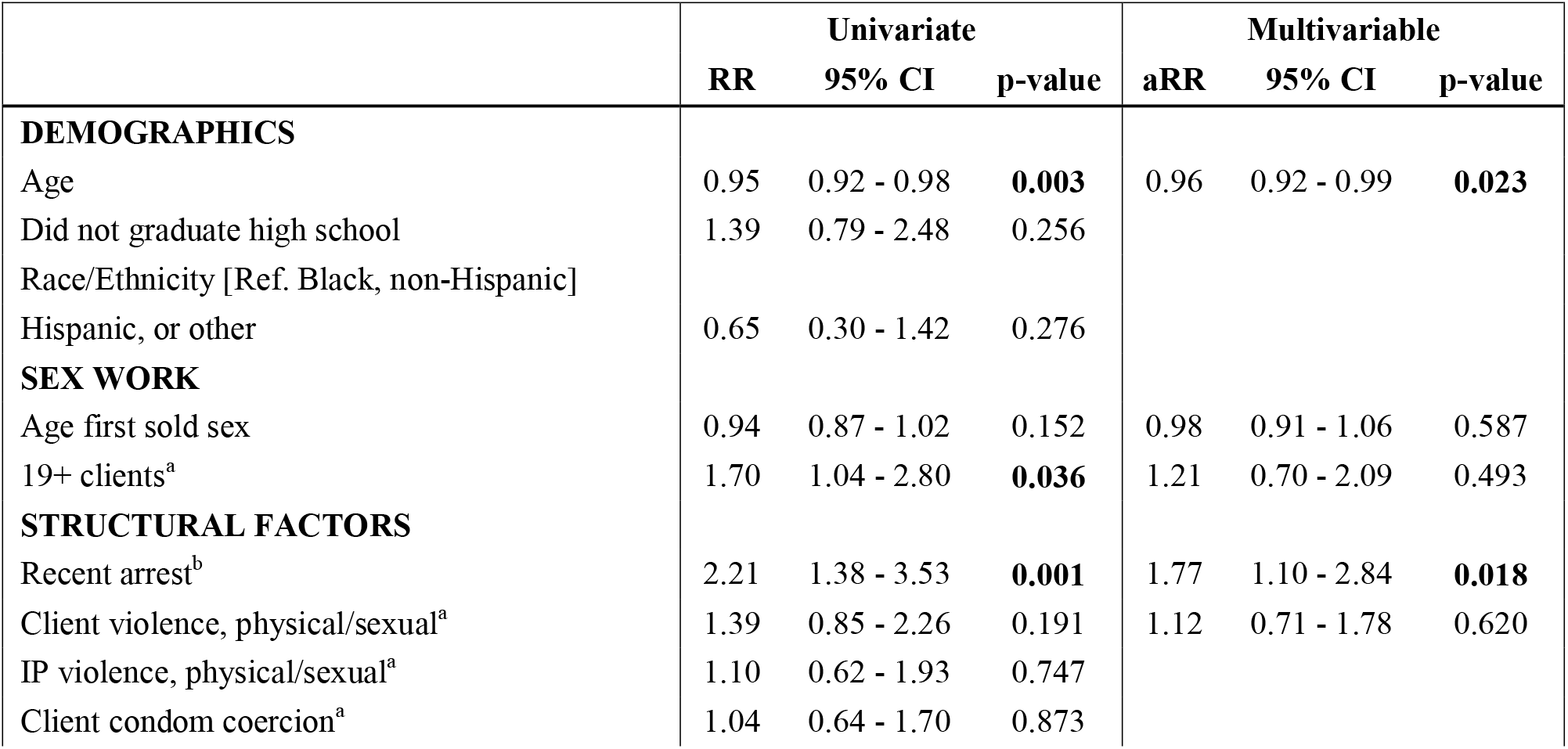
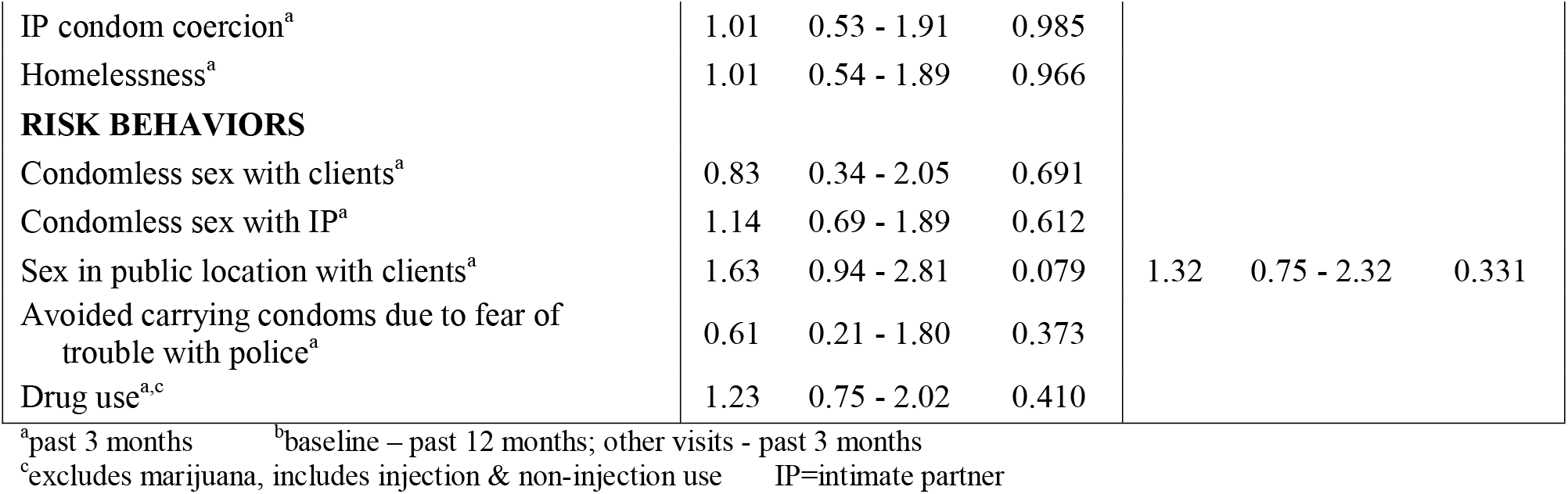
Univariate and multivariable correlates of STI episodes over all study visits using GEE log binomial regressions with exchangeable correlation structure

## RESULTS

### Participant Demographics and Structural Risks

Sixty-two TFSW enrolled in SAPPHIRE and completed the baseline visit. As detailed in Table 1, all TFSW were women of color: 74% (n=46) were non-Hispanic Black and the remaining quarter (n=16) identified as Hispanic, multiracial, or Native American. The mean age was 30 years, and 29% had less than a high school graduate education. Almost one-quarter (24%) experienced homelessness in the past 3 months, the majority were unemployed (79%), yet most had a primary care provider (89%) and reported some form of health insurance (95%). Less than half (37%) had all legal documents that indicated the gender marker they desired. Experiences of physical or sexual violence in the past 3 months by clients and IPs were common (32% and 23%, respectively). Arrest was also common; 81% had ever been arrested and 45% had been arrested in the past year.

### HIV/STI Behavioral Risk Factors

TFSW entered street-based sex work in early adulthood (average age 18 years). At baseline, a third of TFSW reported being high or drunk during sex in the past 3 months; 39% reported condomless anal or vaginal sex with IPs, yet only 8% of TFSW reported condomless anal or vaginal sex with clients in the past 3 months. Receptive anal sex with clients or IPs was more common than insertive anal sex in the past 3 months (81% vs. 53%, respectively). Illicit drug use in the past 3 months, excluding marijuana use, was reported by 44%, and more than half reported binge drinking in the past year (52%). Sex with clients in public locations in the past 3 months was common (86%).

### HIV/STI results

Baseline HIV prevalence was high at 40.3% among TFSW, and only 64% of HIV-positive women were currently on anti-retroviral therapy (Table 1). There were no HIV seroconversions over follow-up. Of the 37 HIV-negative women, 8 (22%) were currently taking Pre-exposure prophylaxis (PrEP) at baseline. The proportion testing positive for NG, CT, and TV at baseline was 9.7%, 17.7%, and 14.5%, respectively (Table 2). More anal specimens than urine samples tested positive for NG, CT and TV at baseline and all follow-up visits, except for TV, for which more urine specimens tested positive at baseline than anal specimens. Only 4 participants had the same STI at the same site at two or more consecutive study visits (data not shown).

**Table 2.** Proportion of positive STI result by specimen source and STI type.

|  | Baseline<br>n=62<br>% | 3-month<br>n=42<br>% | 6-month<br>n=40<br>% | 9-month<br>n=40<br>% | 12-month<br>n=41<br>% |
| --- | --- | --- | --- | --- | --- |
| *Neisseria gonorrhoeae* | 9.7 | 9.5 | 2.6* | 10.0 | 0 |
| Anal only | 6.5 | 9.5 | 2.6* | 10.0 | 0 |
| Urine only | 3.2 | 0 | 0* | 0 | 0 |
| Both anal & urine | 0 | 0 | 0* | 0 | 0 |
| *Chlamydia trachomatis* | 17.7 | 7.1 | 7.5 | 5.0 | 9.8 |
| Anal only | 9.7 | 7.1 | 7.5 | 2.5 | 7.3 |
| Urine only | 6.5 | 0 | 0 | 0 | 2.4 |
| Both anal & urine | 1.6 | 0 | 0 | 0 | 0 |
| *Trichomonas vaginalis* | 14.5 | 14.3 | 10.0 | 10.0 | 9.8 |
| Anal only | 4.8 | 14.3 | 7.5 | 5.0 | 4.9 |
| Urine only | 9.7 | 0 | 0 | 0 | 2.4 |
| Both anal & urine | 0 | 0 | 2.5 | 2.5 | 2.4 |
| Any STI | 35.5 | 23.8 | 15.0 | 20.0 | 17.1 |
| Anal only | 14.5 | 23.8 | 7.5 | 15.0 | 12.2 |
| Urine only | 16.1 | 0 | 0 | 0 | 2.4 |
| Both anal & urine | 1.6 | 0 | 2.5 | 2.5 | 2.4 |

\*n=39

### Univariate and Multivariable regression results

In the univariate regressions for any STI positive result using all study visits, older TFSW were significantly less likely to test positive for an STI (RR 0.95; 95% CI: 0.92-0.98) (Table 3). TFSW with higher client volume were more likely to test positive for an STI across all study visits (RR 1.70; 95% CI: 1.04-2.80), and TFSW who had been arrested recently were also more likely to test positive for an STI (RR 2.21; 95% CI: 1.38-3.53). In the multivariable regression model, both age and recent arrest remained significant. Older TFSW were slightly less likely to experience STIs during study (adjusted risk ratio [aRR] 0.96; 95% CI: 0.92-0.99), and TFSW who were recently arrested were significantly more likely to test positive for an STI (aRR 1.77; 95% CI: 1.10-2.84). Insertive anal sex with clients was associated with increased risk of testing positive via a urine specimen (risk ratio [RR] 3.48; 95% confidence interval [CI]: 1.14-10.62), and we found no statistically significant associations for receptive anal sex by specimen site (Table 4).

**Table 4.** Univariate correlates of any STI at any study visit by specimen site using GEE log binomial regressions with exchangeable correlation structure

|  | Anal Positive |  |  | Urine Positive |  |  |
| --- | --- | --- | --- | --- | --- | --- |
|  | RR | 95% CI | p-value | RR | 95% CI | p-value |
| Receptive anal sex, clients/IP<sup>a</sup> | 1.84 | 0.81 - 4.16 | 0.142 | 1.04 | 0.37 - 2.96 | 0.937 |
| Clients | 1.64 | 0.85 - 3.16 | 0.143 | 1.94 | 0.70 - 5.40 | 0.206 |
| IP | 1.16 | 0.61 - 2.22 | 0.644 | 1.20 | 0.47- 3.09 | 0.705 |
| Insertive anal sex, clients/IP<sup>a</sup> | 1.15 | 0.61 - 2.15 | 0.669 | 2.20 | 0.78 - 6.27 | 0.138 |
| Clients | 1.24 | 0.66 - 2.33 | 0.505 | 3.48 | 1.14 - 10.62 | **0.028** |
| IP | 0.80 | 0.37 - 1.69 | 0.552 | 1.29 | 0.47 - 3.55 | 0.621 |

<sup>a</sup>past 3 months IP=intimate partner

## DISCUSSION

In this longitudinal study with TFSW, we found a high baseline prevalence of HIV and STIs. Unlike Nuttbrock et al.,^8^ we did not see a significant decline in the proportion of STI-positive results over the course of the study. The lack of a reduction in STI-positive results in this study suggests lack of initial treatment and/or ongoing STI exposure. The small number of participants with the same STI at the same site at two or more consecutive study visits is suggestive of new infections rather than lack of treatment. Ensuring that TFSW have access to frequent, gender-affirming STI screening, treatment, and risk reduction interventions is critical to reducing the burden of STIs in this population.

Similar to prior literature,^8^ we found a higher prevalence of condomless sex with IPs than with clients. The low report of condomless sex with clients could be attributable to the outreach efforts of local volunteers who have passed out condoms in areas known to be frequented by TFSW during one night each week for several years prior to data collection for this study. Future research evaluating the efficacy of such outreach interventions for TFSW is warranted.

The association between older age and fewer STIs could be explained by older TFSW having gained more experience in safer sex negotiation strategies. It is also possible that older TFSW have fewer partners. The latter explanation is more likely given that self-reported condom use was not associated with STI results in univariate analysis while number of clients was associated with STIs. Anal STIs (NG, CT, TV) were more common than urethral STIs. However, reporting receptive sex was not significantly associated with having an anal STI; nor was insertive sex associated with having an urethral STI with the exception of insertive sex with clients.

The literature on laboratory-confirmed anal and urethral STIs among transgender women is quite limited.^16^ Prior studies have validated NAAT for NG, CT, and TV in urethral samples^17 18^ as well as NG and CT in anal samples.^19 20^ However, few studies have reported on anal TV,^21-24^ and we found no studies of anal TV in transgender women. Reported prevalence of anal TV among MSM have ranged from <1%^20 24^ to 9%.^21^ Similarly, prevalence of anal TV among cisgender women has ranged from 1.2%^22^ to 6.3%.^23^ Our finding of positive anal TV results ranging from 4.8% to 14.3% across study visits is consistent with the studies in cisgender populations. Few participants tested positive for both anal and urethral TV. This is also consistent with prior studies and suggests that anal TV is unlikely to be a contaminant from the urethra. More research is needed to understand the role of anal TV in the burden of STIs among transgender women, particularly TFSW who are highly vulnerable to STIs.

Prior studies have demonstrated the value of STI screening of multiple anatomic sites.^25^ Recent data specifically highlight the proportion of STIs missed when anal testing is not done. Jordan et al. evaluated 3,191 STI testing kits from a national internet-based screening program; among females and males, 17.1% and 16.7% of TV positive samples were missed, respectively.^26^ In our study, 32 anal STIs would have been missed if only urethral testing was done. Our finding of no correlation between reported sexual positioning and site of STI highlights the importance of testing at all sites of potential exposure, regardless of self-reported sexual positioning.

In the baseline univariate analyses, race and education were significantly associated with testing positive for an STI. Such racial and socioeconomic disparities in STI prevalence have been well documented.^5 16^ The confluence of racism, classism, transphobia, and criminalization in the lives of TFSW result in heightened vulnerability to negative health outcomes, including STIs.^4^

In multivariable analyses, recent arrest was the only structural factor that remained significantly associated with positive STI results – both baseline STIs and STIs over the course of the study. Street-based TFSW of color are particularly vulnerable to arrest. They are often profiled by police as sex workers simply because they are “walking while trans.”^27^ Arrest results in increased economic pressure caused by costs associated with money bail and court fees as well as loss of income while incarcerated. These economic pressures may in turn lead TFSW to increase the number of sex work clients in order to mitigate these economic losses. As illustrated by the baseline analyses, we found that TFSW with an STI had a significantly higher number of sex partners than women who tested negative for STIs. Therefore, reducing arrest may be an important way to reduce STI risk among TFSW.

Sexual violence against transgender women is common in jails and prisons in the U.S.^4 28^ When incarcerated, transgender women are frequently housed with cisgender men, putting them at high risk for violence.^28^ Research indicates that decriminalization of sex work could significantly reduce the incidence of HIV, in particular, among sex workers.^29^ Advocates for the health and human rights of sex workers have long called for decriminalization of sex work and the implementation of workplace protections.^30^ Our data support this recommendation.

The small sample size and high rate of attrition over the course of the study reduced the statistical power to detect significant differences. Self-reported HIV risk behaviors, such as condom use, could not be verified, therefore were subject to recall and social desirability bias. This study tested for only a subset of all bacterial STIs and lacked data on oral sex and oropharyngeal STI testing. However, the study was strengthened by the use of laboratory-confirmed measures of both HIV and STI outcomes over time, including assessments at urethral and anal sites of exposure and multiple types of STIs. Compared to both CFSW and transgender women who do not engage in sex work, TFSW bear a heavy burden of both HIV and STIs.^10^ Future research is needed with larger sample sizes and to allow for more complex analyses. Future studies should include a broader range of common STIs such as syphilis, herpes simplex virus, and human papillomavirus and test multiple potential sites of exposure, including oropharyngeal sites.

## Data Availability

De-identified participant data are available from Dr. Susan Sherman at Johns Hopkins University: https://orcid.org/0000-0001-8399-7544 upon reasonable request and review after submission of a manuscript concept form.

